# Structured tools to assessing quality and bias in Mendelian randomisation studies: an updated systematic review

**DOI:** 10.1101/2025.09.05.25335166

**Authors:** Jinyue Yu, Mengxuan Zou, Francesca Spiga, Sarah Dawson, George Davey Smith, Julian PT Higgins

**Author notes:** Corresponding Author: Professor Julian PT Higgins Bristol Medical School (Population Health Science) University of Bristol.

## Abstract

**Background:** The growing use of Mendelian randomisation (MR) has heightened the need for rigorous quality and bias assessment tools. A previous systematic review included studies published up to July 2021 identified 14 structured instruments for conducting, evaluating, and reporting MR studies. However, methodological developments have accelerated in the years since.

**Methods:** We updated Spiga’s systematic review to include tools published between July 2021 and January 2025, applying the same search strategy and eligibility criteria. Two reviewers independently screened articles, extracted data, and mapped tool content to bias domains.

**Results:** We identified 15 additional articles, bringing the total to 29 tools. Of these, 19 provided structured evaluation tools. 12 of the 19 evaluation tools were newly added in the present review, which addressed broader methodological domains beyond core instrumental variable assumptions, including genetic instrument selection, population stratification, sensitivity analyses, and dataset considerations. However, substantial variation in bias domains, structure, and scoring methods across tools persists. Key gaps remain in the assessment of linkage disequilibrium, missing data, and dynastic effects.

**Conclusions:** While the number of structured tools has increased in recent years, the lack of standardisation across tools still makes it difficult to assess the reliability and comparability of MR studies. Developing more complete and standardised evaluation frameworks and properly testing these tools in practice are important next steps to improve the overall quality of MR research.

**Key Messages:**

- Mendelian randomisation (MR) studies are increasingly applied, but there is still no standard approach for assessing their quality and risk of bias.
- Our updated systematic review identified 15 new articles since the review by Spiga et al., which included studies published up to July 2021, bringing the total to 29 tools, with 19 providing structured tools for evaluation.
- Newer tools cover a broader range of methodological domains beyond the three core instrumental variable assumptions, including genetic instrument selection, population stratification, sensitivity analyses, and dataset considerations.
- There are still large differences between tools in structure, scoring methods, and bias domains covered, and some important areas such as linkage disequilibrium and missing data remain under-assessed.

## Introduction

Mendelian randomisation (MR) has become a highly-used approach in epidemiological research, offering a robust method for causal inference, generally through leveraging genetic variants as instrumental variables (1-4). The appeal of MR lies in its ability to reduce confounding and reverse causation, common limitations in conventional observational studies, making it a valuable tool for strengthening causal inference (3, 5).

With the rapid expansion of MR applications across various domains of biomedical research, there has been an accompanying growth in concern regarding the robustness, reproducibility, and interpretability of many published MR studies. Editorials and commentaries have raised alarm over the uncritical or superficial use of MR methods, prompting calls for more rigorous and standardised approaches to evaluating study quality and potential biases (4, 6-8). The complexity of MR studies, including the selection of appropriate genetic instruments, assumptions underpinning instrumental variable analysis, and sensitivity to population structure, necessitates structured tools to guide researchers in evaluating methodological robustness (8, 9). Over the years, several tools have been developed to aid in the appraisal of MR studies, providing structured frameworks for evaluating risk of bias, assessing validity, and ensuring methodological consistency (10). Despite these efforts, no universally accepted, standardised approach to MR risk of bias and quality assessment has been established.

Variability in assessment criteria and inconsistency in tool usage across studies highlight the need for continued refinement and harmonisation of evaluation methods. Addressing this gap is essential to enhance the credibility and reproducibility of MR findings, particularly in the context of systematic reviews and meta-analyses.

A previous systematic review identified and evaluated tools for assessing risk of bias and study quality in MR research published up until the first half of 2021 (10). The review by Spiga et al. identified 14 tools designed for the evaluation, conduct, and/or reporting of MR studies, with seven tools specifically developed for risk-of-bias or quality assessment. These tools addressed key MR-specific biases, particularly those related to instrumental variable assumptions, genetic instrument selection, and population/sample characteristics. While this review provided an essential foundation for MR assessment, further investigation was needed to identify additional tools that have emerged since 2021 following the publication of STROBE-MR (8) and to evaluate the value of these tools in supporting the conduct, evaluation, and reporting of MR studies in systematic reviews.

Building on this earlier work, we updated the systematic review to identify newly developed tools, or tools adapted from existing frameworks, for assessing MR studies. This update aims to synthesise recent developments on MR assessment tools systematically, providing a comprehensive and current overview of available methodologies for considering the quality of MR studies.

## Methods

### Eligibility Criteria

We retained the eligibility criteria used by Spiga et al. (2023) (PROSPERO (CRD42021282836) though restricted our update to newly developed or significantly modified structured tools for evaluating MR conduct, evaluation and/or reporting (Spiga et al.’s “review of tools” rather than their “review of systematic reviews”). Eligible articles needed to (i) present structured tools—such as guidelines, checklists, or frameworks— designed to support comprehensive assessment of the design, conduct, and/or reporting of Mendelian randomisation (MR) studies, or to provide systematic guidance through the process of conducting or reporting such studies; and (ii) be available as full-text articles, including peer-reviewed publications, preprints, or published protocols published since July 2021. We excluded articles that merely referenced existing tools (e.g., STROBE-MR) without substantial modification, or that did not employ a structured evaluation method.

### Search Strategy

We systematically searched MEDLINE (Ovid), Embase (Ovid), Web of Science, preprint servers (bioRxiv and medRxiv), PROSPERO, Google Scholar for articles published since Spiga et al’s search in July 2021 up to 24 January 2025. We adopted the same search strategy as Spiga et al. for MR tools (see supplementary materials). Moreover, we reviewed the aims and scope of all 45 protocols listed in Spiga et al (10), searching publication databases to identify any subsequent full-text publications arising from the protocols. No language restrictions were imposed. References from identified articles were screened for additional eligible articles.

### Study Selection and Data Extraction

Two reviewers (JY and MZ) independently screened titles and abstracts for relevance using Rayyan app (www.rayyan.ai). Full texts of potentially eligible articles were independently assessed by the same reviewers, with discrepancies resolved through discussion or by consulting a third reviewer (FS).

For each included study, we extracted details about: the type of tool (newly developed or modified); specific IV assumptions assessed; criteria used for assessment and any scoring system applied; and practical application of the tool in MR research or systematic reviews.

### Data Synthesis

We summarised findings in a narrative synthesis, describing the characteristics of new or modified MR risk-of-bias tools. To ensure consistency and comparability with the original review by Spiga et al (10), we adopted their methodological approach for categorising assessment items within each identified tool. We systematically reviewed each structured evaluation tool included in our update, extracting individual assessment items. These items were then categorised according to the specific type of bias or methodological concern they addressed, and subsequently grouped into clearly defined bias-related domains.

In line with the original review, we anticipated that most tools would fall into one of three previously established categories: i) tools designed to support the conduct of MR studies, ii) tools to assess risk of bias or study quality, and iii) tools to guide reporting. Where any newly identified tools did not fit these categories, we classified them under a new category, if appropriate, to reflect their distinct purpose.

As this review synthesises methodological tools rather than intervention or outcome data, a formal assessment of certainty or reporting bias was not applicable

## Results

### Summary of the screening

As shown in Figure 1, our updated literature search identified a total of 1112 records. We additionally identified seven potentially relevant articles arising from the 45 protocols listed in Spiga et al. (2023). After deduplication, 646 unique articles underwent title and abstract screening, and 36 full-text articles were assessed for eligibility of inclusion in the review. Of these, 15 new tools met our inclusion criteria and were included in the current update, two of which (11, 12) were identified from the afore-mentioned protocols. Combining these with the 14 tools from the original review by Spiga et al. (2023), a total of 29 tools were included in this systematic review.

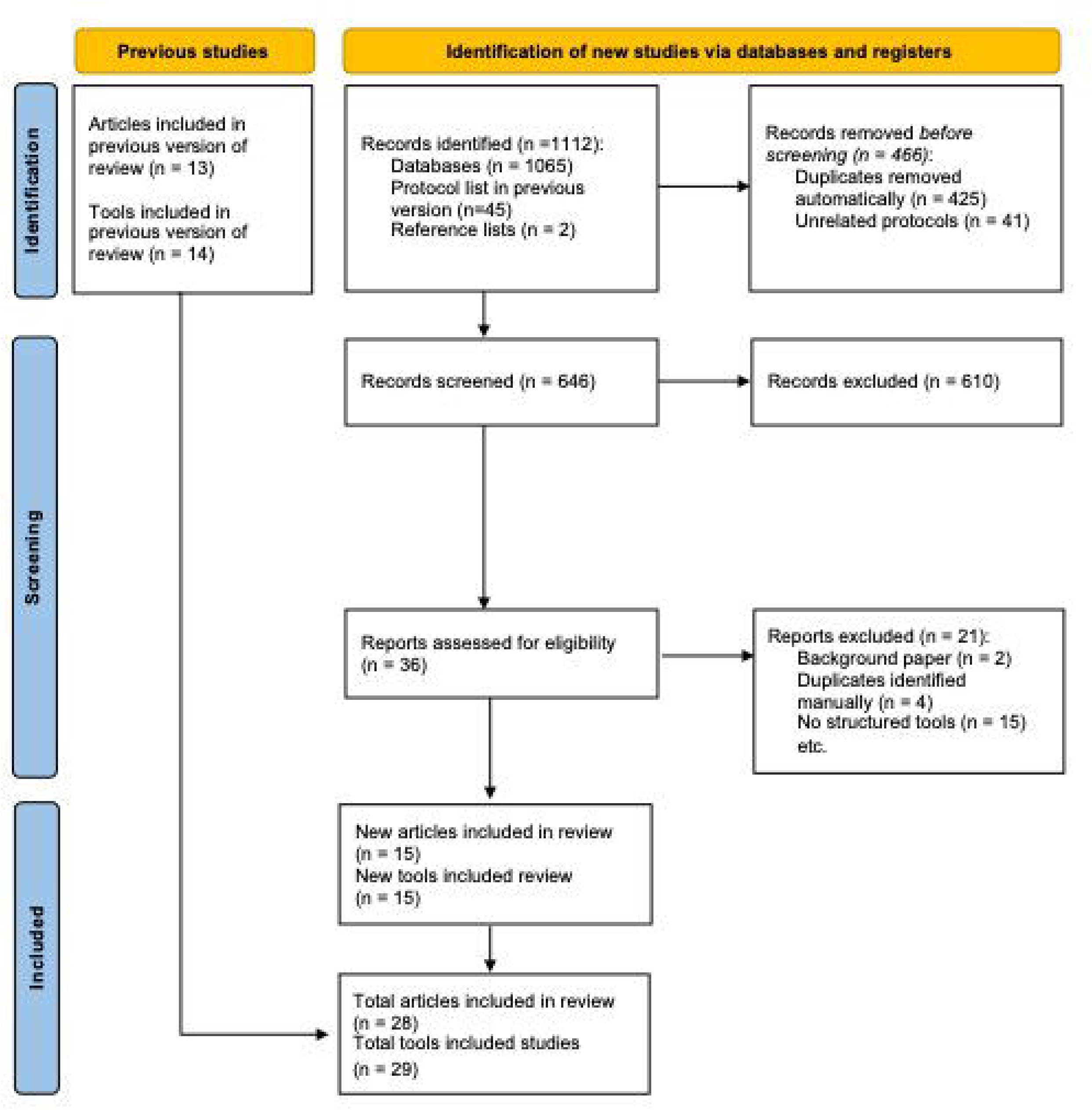

Table 1 summarises the scope of the 29 included tools. A total of 19 were structured tools specifically designed for assessing methodological quality or risk of bias in MR studies (7 had been reported in Spiga’s review, 12 were newly identified in this update). These tools included assessments of the three IV assumptions (13-16), domain-based checklists or rating scales (17-23), and novel scoring metrics (24, 25). Moreover, a total of 13 tools included assessment of reporting quality of the MR study; five tools explicitly focused on structured tools aimed at improving reporting quality in MR research, including checklists specifically for MR studies (8, 20, 26-28); eight tools addressed both reporting and evaluating purposes within one tool. Additionally, four tools provided structured guidance explicitly designed for conducting MR studies (18, 29-31), and one tool identified in this update (12) was classified into a new category as a tool for assessing “robustness of the evidence”. Burgess et al, (18) uniquely addressed all three purposes—evaluating, conducting, and reporting in two separate tools in 2020, and then substantially revised and published the guidelines in 2023 (7). The updated version retains the original two-part structure (a checklist and a flowchart) but has significantly expanded its content and scope. It now includes ten thematic sections covering motivation, data sources, variant selection, harmonisation, primary and sensitivity analyses (including robust methods), data presentation, and interpretation. Moreover, the revised version introduces new guidance on within-family MR to address biases such as dynastic effects and assortative mating, offers updated recommendations for drug-target MR, and provides a detailed summary table of robust MR methods.

**Table 1.** Details of included studies containing one or more tools for evaluating, conducting, and reporting MR studies.

| Tools | Study ID | Type of tool | Scope of the tool(s) | Number of tools | Structure of the tool |
| --- | --- | --- | --- | --- | --- |
| 1 | Boef 2015 | Systematic review of methodological approaches and quality of reporting in MR studies | Reporting | 1 | Checklist |
| 2&3 | Burgess 2020 | Guideline for performing MR investigations | Assessing * | 2 | 1. Domain-based checklist (assessing and reporting)<br>2. Flowchart (conducting) |
|  |  |  | Conducting |  |  |
|  |  |  | Reporting |  |  |
| 4 | Davey Smith 2019 | STROBE-MR; guideline for the reporting of MR studies | Reporting | 1 | Checklist |
| 5 | Davies 2018 | Guide, checklist and glossary for MR studies | Assessing | 1 | Checklist |
|  |  |  | Reporting |  |  |
| 6 | Grau-Perez 2019 | Systematic review of MR studies | Assessing | 1 | Domain-based evaluation chart |
|  |  |  | Reporting |  |  |
| 7 | Grover 2017 | Guideline for performing MR analysis | Conducting | 1 | Flowchart |
| 8 | Kuźma 2018 | Systematic review of MR studies | Assessing | 1 | Rating scale |
|  |  |  | Reporting |  |  |
| 9 | Lawlor 2019 | MR dictionary and online tool | Conducting | 1 | Flowchart |
| 10 | Lee 2020 | Protocol for a systematic review of MR studies on risk factors for dementia | Assessing | 1 | Checklist |
|  |  |  | Reporting |  |  |
| 11 | Lor 2019 | Systematic review of MR studies and guideline for reporting MR studies | Reporting | 1 | Checklist |
| 12 | Mamluk 2020 | Systematic review of MR studies | Assessing | 1 | Bias domain-based rating |
| 13 | Swerdlow 2016 | Review of selection of instrument in MR analysis and systematic review of MR studies | Conducting | 1 | Decision tree |
| 14 | Treur 2021 | Systematic review of MR studies | Assessing | 1 | Scoring system |
|  |  |  | Reporting |  |  |
| Updates 2021-2025 |  |  |  |  |  |
| 15 | Bouajila 2024 | Systematic review of MR studies | Assessing | 1 | Domain based rating |
| 16 | Burgess 2023 | Checklists for assessing MR studies | Assessing | 1 | Checklist |
| 17 | Luo QF 2024 | Review of MR studies in osteoarthritis risk | Assessing | 1 | Assumption-based rating |
| 18 | Luo S 2022 | Systematic review of MR studies | Assessing | 1 | Assumption-based rating |
| 19 | Omar 2024 | Systematic review of MR studies | Assessing | 1 | Assumption-based rating |
| 20 | Nguyen 2024 | Guide for MR studies | Assessing | 1 | Checklist |
|  |  |  | Reporting | 1 |  |
| 21 | Skrivankova 2021 | STROBE-MR; guideline for the reporting of MR studies | Reporting | 1 | Checklist |
| 22 | Woolf 2022 | Guideline for the reporting of 2SMR studies | Reporting | 1 | Checklist |
| 23 | Gao 2024 | Systematic review of MR studies | Assessing | 1 | Domain-based rating |
| 24 | Gibson 2023 | Systematic review of MR studies | Assessing | 1 | 25-item checklist |
|  |  |  | Reporting |  |  |
| 25 | Alhassan 2024 | Scoping review of MR studies | Assessing | 1 | Assumption-based rating |
| 26 | Visontay 2022 | Systematic review of MR studies | Assessing | 1 | Bias domain-based rating |
| 27 | Rosas-Chavez 2025 | A metric for quality evaluation of MR studies | Assessing | 1 | Scoring system |
| 28 | Lee 2024 | Systematic review of MR studies | Assessing | 1 | Bias domain-based rating |
| 29 | Markozannes 2022 | Systematic review of MR studies | Robustness of the evidence | 1 | Grading flowchart/framework |

Notes: Abbreviations: MR=Mendelian randomization; STROBE= Strengthening the Reporting of Observational Studies in Epidemiology; 2SMR= Two-Sample
Mendelian Randomisation; \*Assessing =assessing risk of bias/quality of evidence.

### Tools for evaluating quality/risk of bias of the MR study

The bias-related domains addressed by the 19 tools for evaluating methodological quality or risk of bias are summarised in Table 2. A total of 150 items addressed biases/quality related issues, 77 of which directly related to the three core IV assumptions. All tools explicitly addressed the independence assumption (IV3), emphasising consistent attention towards horizontal pleiotropy. The independence assumption (IV2), particularly in relation to confounding, was addressed by the majority of tools (84%). The relevance assumption (IV1), with a focus on weak instrument bias, was also assessed by most tools (89%). Frequently evaluated additional domains included sensitivity analyses (74%), sample overlap (42%), and population heterogeneity (42%). Less commonly addressed domains included choice of controls (5%), construction of genetic score (5%), and missing data (11%).

**Table 2.**
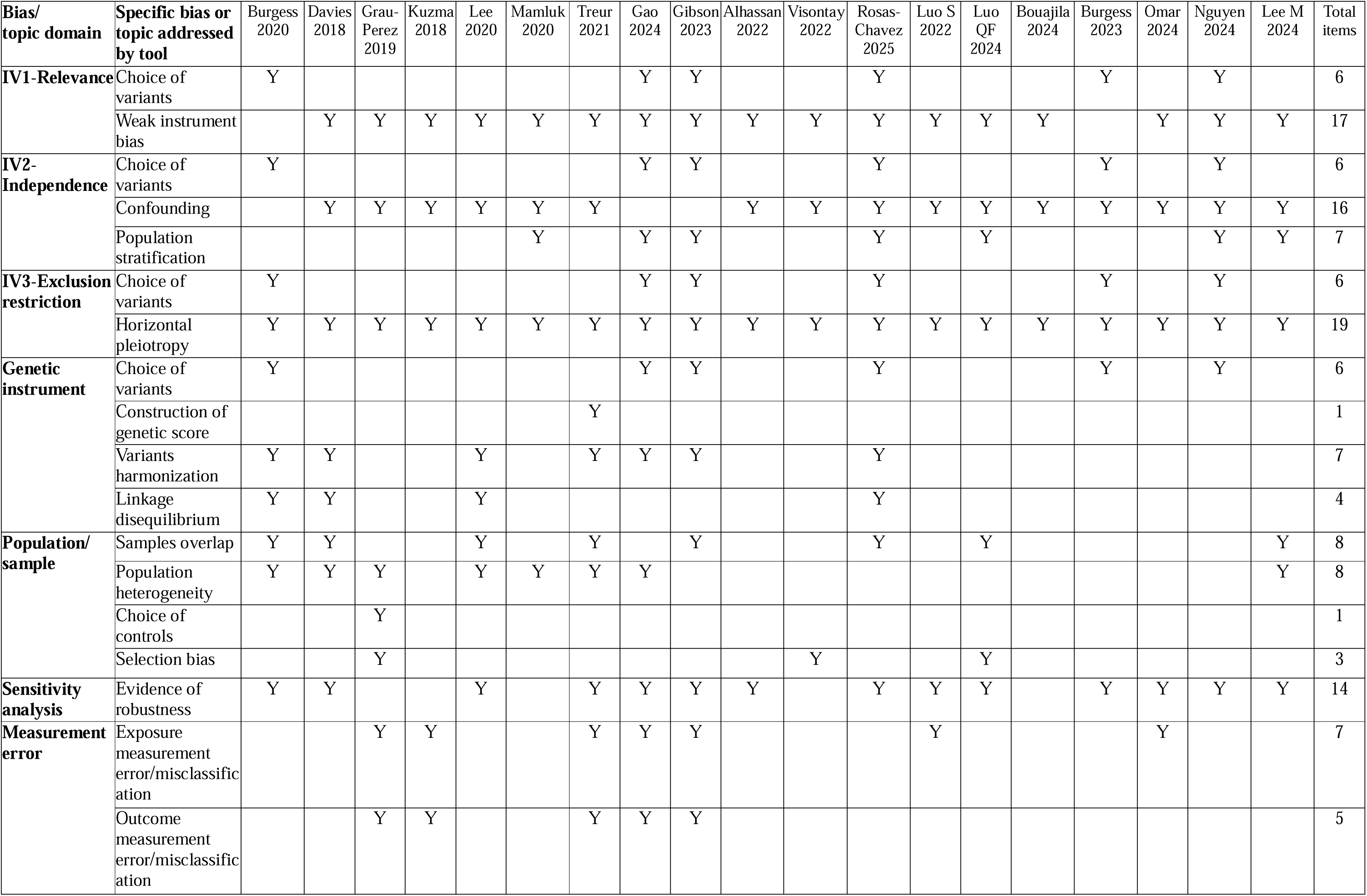
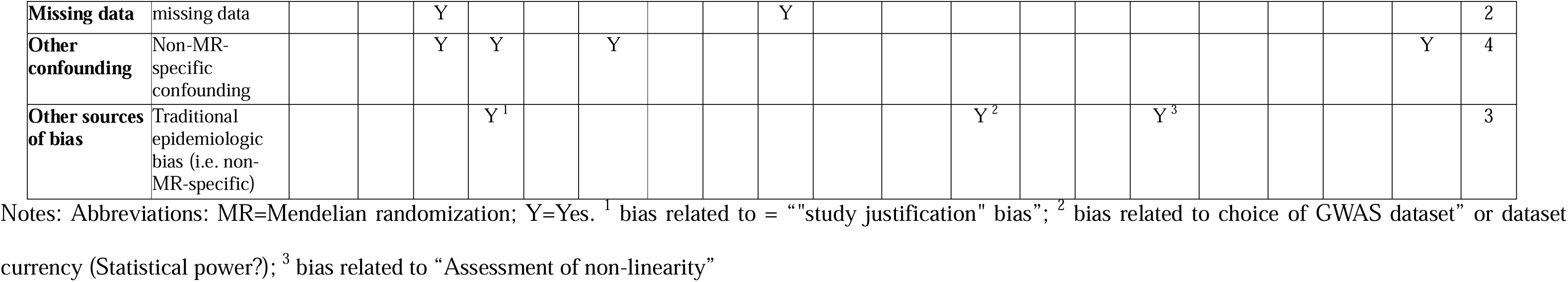
Details of specific Mendelian randomization bias and limitation addressed by items or questions within each assessing tool.

Additionally, three tools assessed additional biases not explicitly listed in the standard domains. Specifically, Kuzma (2018) (32) addressed bias related to "study justification"; Rosas-Chavez (2025) (24) included an assessment of statistical power and choice or currency of GWAS datasets; and Bouajila (2024) (17) introduced assessment of non-linearity in their evaluation system.

### Tools for addressing conducting and reporting quality of MR study

Among the 13 reporting tools, the number of domains ranged from 3 to 6, with all tools addressing reporting of the three core IV assumptions. Woolf et al (28) included items addressing homogeneity and sample overlap (specifically in two-sample MR studies), while Gibson et al (20) addressed linkage disequilibrium and heteroscedasticity. For tools guiding conduct of MR studies, Grover et al (29), Lawlor et al (30), and Swerdlow et al (31) covered between 5 and 10 domains, including items addressing the selection of genetic instruments, assumptions validation, handling of pleiotropy, and statistical analyses.

### A new tool for assessing robustness of the evidence from an MR study

We identified a newly developed framework that focuses on evaluating the strength and consistency of MR findings; Markozannes et al. (12) proposed a structured decision flow that classifies each exposure-outcome association into one of five levels: non-evaluable, insufficient, suggestive, probable, or robust evidence. This classification depends on whether the main MR estimate reaches statistical significance and whether results from additional methods, such as MR-Egger, weighted median, MR-PRESSO, or multivariable MR, show consistent direction and significance. Although many of the included tools addressed “evidence of robustness” as part of their risk-of-bias assessment, this generally referred to whether sensitivity analyses were performed, such as MR-Egger or MR-PRESSO. These items did not assess how consistent or supportive the findings were across different methods. Therefore, we created a new category to reflect this distinction, which may merit further research attention.

### Other aspects for consideration of MR study

We summarised details of 57 items addressing any other aspects of the MR analysis in Table 3. Among these, we found two items in two tools addressing clinical implications of MR results (33, 34); six items in six tools addressing the datasets used (7, 20, 24, 33-35); 35 items in 17 tools addressing the genetic instrument, including genetic variant selection rationale (7, 14-20, 23, 24, 34), method used to obtain variants (7, 13-17, 19-21, 23, 24, 34), and variant strength (12-17, 19, 20, 23-25, 34); 23 items in 10 tools addressing the interpretation of MR analysis results, including applicability, transportability, temporality, and bidirectional effects (7, 14, 17, 19, 20, 23-25, 33, 34); 12 items in 12 tools addressing the MR rationale (7, 14-18, 20, 23-25, 32, 34); 19 items in 15 tools addressing the MR results (7, 12-17, 19-21, 23, 24, 33-35); five items in five tools addressing precision of the results, including statistical power or sample size (21, 24, 25, 32, 34); eight items in five tools addressing the selection of the population(s) or sample(s) (12, 14, 18, 20, 24); and 36 items in 12 tools addressing statistical analysis methods and any related details including reporting of primary and secondary analyses (7, 12-14, 18-21, 24, 25, 32, 34); and five item in five tools addressed the type of dataset utilised (7, 18, 20, 24, 34).

**Table 3.**
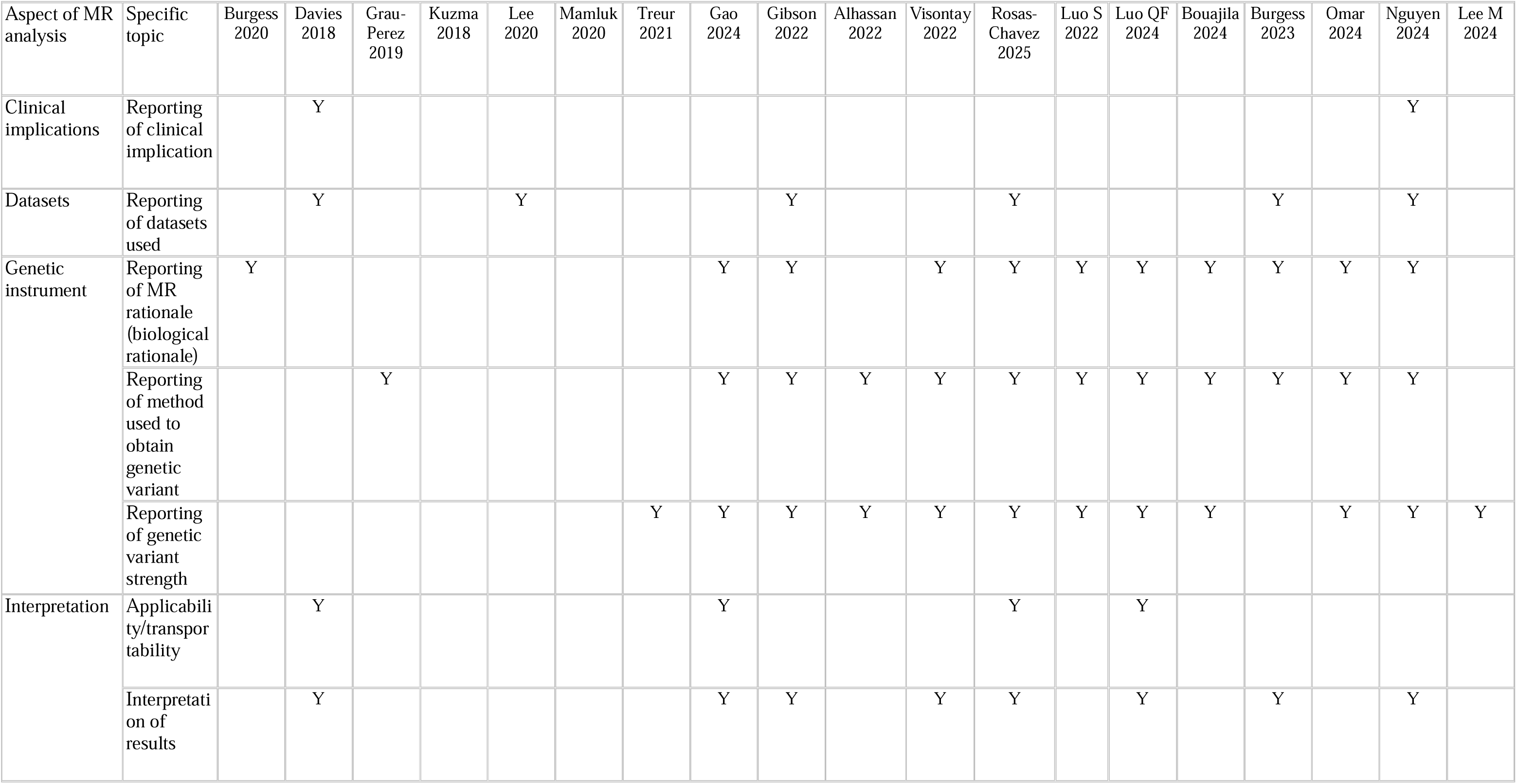
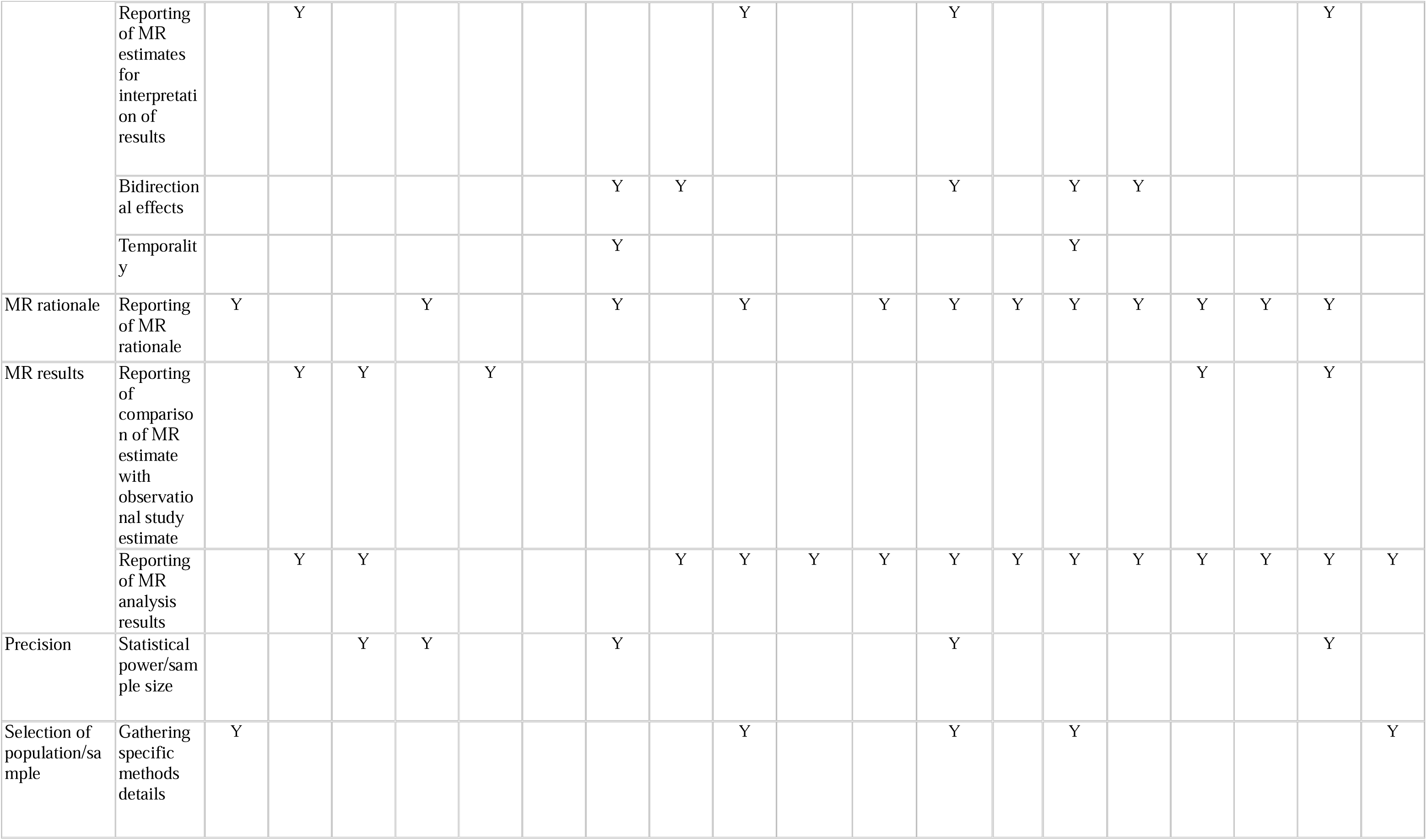
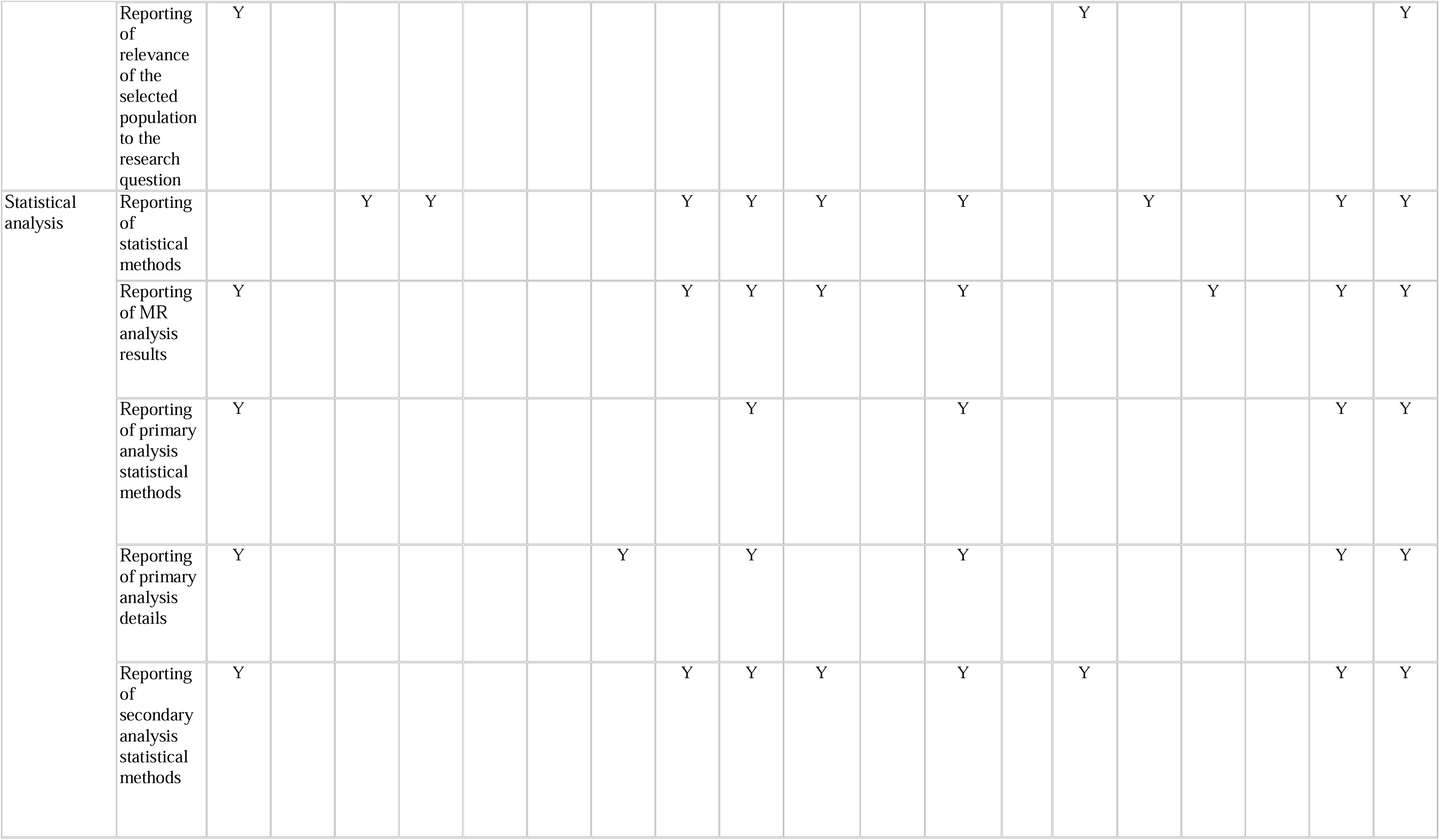
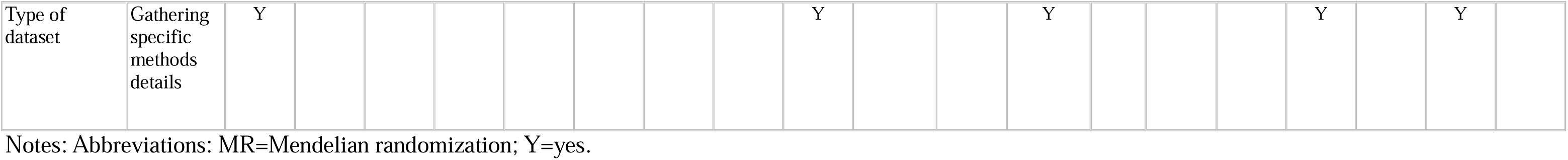
Details of other MR-relevant content of items or questions within each assessing tool.

## Discussion

In this updated systematic review, we identified 12 new structured tools for assessing the quality and risk of bias in MR studies. This doubles the number of tools identified by Spiga et al (10). Of the total 29 tools included, 19 provided structured tools mainly focused on evaluating MR quality and bias. The newer tools not only continue to assess the three key IV assumptions but also broaden their scope to cover areas such as genetic instrument selection, population stratification, sensitivity analyses, and other important methodological aspects.

Moreover, we observed differences in how these domains are assessed by different tools, including bias-domain based rating, assumption-based rating, scoring systems, and checklist.

Since publication of the earlier review, our update shows a clear trend of increase in the number of structured evaluation tools and an increase of MR in systematic review, which need improved assessment approaches. This reflects the growing awareness among researchers of the need to assess systematically the quality and potential biases in MR studies. Importantly, our detailed assessment revealed that newly developed tools generally include more extensive and varied items across assessment domains compared with earlier tools; as shown in Table 3, newer tools tend to go beyond the basic IV assumptions, and consider additional factors such as robustness through sensitivity analyses. This development suggests a positive step towards creating more comprehensive frameworks for evaluating MR studies; however, the inclusion of these additional aspects remains limited and inconsistent across tools.

Notably, we identified a new category of assessment tool in this update. The robustness of evidence tool by Markozannes et al. (12) provided a structured framework to evaluate the consistency and strength of MR findings across multiple analytical methods. This approach differs from conventional tools that assess study conduct, internal validity, or reporting quality. In contrast, it aims to support interpretation of results by classifying each MR association into levels of evidence, depending on agreement across various robustness checks. While many risk-of-bias tools assessed whether MR studies conducted appropriate sensitivity analyses to address aspects of robustness(7, 11, 13-16, 18-20, 24, 25, 33-35), we see the more holistic assessment of “evidence of robustness” presented by Markozannes et al as a distinct purpose. Such an assessment is not designed to address bias directly, and it may complement risk-of-bias tools by helping researchers judge whether a causal interpretation is supported consistently across analyses. As this was the only application of such tools we found, future consideration might be made as to whether formal assessment of result robustness should become a standard component of MR appraisal, particularly when findings are used to guide causal inference or policy recommendations.

Another development since the Spiga review is the substantial update to the Burgess et al. guidelines, published as a revised version in 2023 (7). While the earlier version had already provided a dual-tool structure for evaluation and conduct, the updated guidelines now encompass a more comprehensive and methodologically detailed framework. Key additions include new sections on within-family MR, updated recommendations for drug-target analyses and variant selection, and expanded coverage of robust statistical methods and triangulation strategies. The inclusion of visual tools such as flowcharts, a structured reviewer checklist, and a comparative table of MR methods further enhance its practical utility. These enhancements reflect ongoing concerns about methodological rigour in MR studies and signal a shift towards more formalised approaches to quality and bias assessment.

Nevertheless, despite the wider coverage of biases, some challenges remain in current evaluation tools. One key issue is the lack of standardisation. Different tools use different wording and different structures (e.g. checklists, ratings, and scoring systems), which would make it difficult to compare results across them. Furthermore, while the three core IV assumptions are widely covered, other important issues, such as linkage disequilibrium, missing data, and dynastic effects, are still not addressed consistently.

Moreover, a fundamental but often underappreciated assumption in MR is gene-environment equivalence (36, 37), which requires that genetic proxies for exposures exert effects that mimic those of modifying the exposures through feasible means. This concept, closely linked to the consistency assumption in causal inference (38), has already been noted in STROBE-MR (8, 9) and the previous Spiga review (10). Failure to meet this assumption, particularly when the genetic instruments are associated with traits in ways that differ mechanistically from the ways in which exposures would be modified through any actual intervention, can lead to biased or misleading inferences. This issue is increasingly recognised as a central problem in MR, potentially exceeding concerns about weak instruments or horizontal pleiotropy (39). Our updated review highlights that, although this assumption is conceptually critical, it is only mentioned by a few tools (7, 11, 14, 34), and among these, only Nguyen et al. (34) included it as an independent checklist item. This represents a methodological gap, particularly where exposure mechanisms differ between genetic variation and potential interventions. Recent publications have highlighted these and other problems in MR practice, including poor variant selection, superficial sensitivity analyses, and overinterpretation of findings (3, 39). Our review supports these concerns, revealing continued inconsistency in how key sources of bias are evaluated.

Another trend identified by our update was a reduced emphasis on reporting-specific tools since the establishment of the STROBE-MR guidelines in 2021. The maturity of STROBE-MR likely accounts for this shift, as it provides a widely accepted framework for enhancing transparency in reporting. Conversely, we observed a substantial rise in tools explicitly developed to evaluate methodological quality and risk of bias, indicating a current gap and ongoing debate regarding optimal assessment methodologies in MR studies. Moreover, our findings indicate that tools designed explicitly for guiding the conduct of MR studies remain limited. The relatively low number of conducting-specific tools suggests an under-explored area, highlighting a critical direction for future methodological research to support robust study design and implementation.

The strengths of this updated review include its timely response to the rapid expansion of methodological research in MR, doubling the number of included evaluation tools compared with the previous review. While this represents important progress, it remains modest in the context of the exponential growth in published MR studies in recent years (4, 6).

Nevertheless, the findings from this review provides substantial evidence and comprehensive reference material essential for developing future standardised MR assessment frameworks. We ensured complete consistency with the original systematic review in search strategies and inclusion criteria, strengthening the reliability of our systematic approach. Importantly, we identified and added a new category that covers tools designed to assess the robustness of the evidence from MR analyses. We also highlighted several types of “ other sources of bias” that have not been categorised in any existing domain/bias topics, such as statistical power (24), study justification (32), and assessment of non-linearity (17), which may warrant integration into future standardised MR evaluation frameworks. Another strength of this review is its focus on tools published after 2021—the year when the STROBE-MR reporting guideline was introduced. The availability of a standardised reporting framework has led to increased awareness of methodological quality in MR studies. As a result, researchers have begun shifting attention from reporting transparency to the more complex and unresolved area of risk-of-bias assessment. This shift is reflected in the increase in newly developed tools since 2021, with our review identifying 12 new structured evaluation tools. By consolidating these developments, we hope to accelerate progress toward the creation of a robust and standardised tool for bias assessment in MR studies, which can support their appropriate use as part of a triangulation of evidence approach in epidemiological research (4, 6, 40).

We also acknowledge limitations in the present review. Our classification of items into different types of biases was subjective and others may reach different conclusions. Furthermore, we searched key sources of grey literature, including preprint servers and registered protocols, but did not include other sources such as conference proceedings, theses, or institutional repositories. Therefore, it is possible that tools could have been missed by our search strategy.

As highlighted in our review and recent publications (4, 6, 41), concerns persist regarding the quality and interpretability of many MR studies. These include the superficial application of methods, poor instrument selection, insufficient sensitivity analyses, and limited attention to the assumption of gene-environment equivalence, as discussed above. These criticisms reinforce the need not only for structured assessment tools but also for greater adoption of these tools to improve transparency, reproducibility, and methodological rigour.

This updated review reflects recent methodological progress in MR study evaluation, highlighting remaining challenges, particularly around standardisation of bias assessment criteria. Future research should aim to address these gaps, promote consensus on methodological standards, and consider the relevance of additional bias domains identified here, ultimately improving the quality, transparency, and reproducibility of MR studies.

Moreover, given the volume of MR studies and persistent concerns about quality, future studies should aim for standardised and potentially semi-automated assessment tools. Such tools could help improve rigour and reproducibility while reducing the burden on peer reviewers and editors.

## Data Availability

All data produced in the present work are contained in the manuscript

## Declarations

### Ethics approval and consent to participate

Not applicable.

### Consent for publication

Not applicable.

## Author Contributions

JPTH and FS conceived the study. SD designed the search strategy. JY and MZ screened the search results and extracted data. JY performed the synthesis and drafted the manuscript. JPTH, GDS, FS, SD, and MZ critically revised the manuscript. JPTH provided overall supervision. All authors reviewed and approved the final version of the manuscript.

