## Supplemental materials for "Structured tools to assessing quality and bias in Mendelian randomisation studies: an updated systematic review"

### Supplementary data

**Search strategy for tools aimed at evaluation of the design, conduct and/or reporting of Mendelian randomization studies**

24 Jan 2025

**Ovid MEDLINE(R) <1946 to present>**

**Embase <1974 to 2025 January 24>**

1. Mendelian randomi#ation.hw,tw,kw,kf.

2. genetic instrument?.hw,tw,kw,kf.

3. (genetic variant* adj3 instrumental variable*).tw,kw,kf.

4. or/1-3

5. scale*. tw,kw,kf.

6. checklist*. tw,kw,kf.

7. critical apprais*.tw,kw,kf.

8. tool*. tw,kw,kf.

9. guide*. tw,kw,kf.

10. (Dictionary or glossary). tw,kw,kf.

11. or/5-10

12. valid*. hw,tw,kw,kf.

13. quality. hw,tw,kw,kf.

14. ((bias OR confound*) AND (asses* OR measur* OR evaluat*)).tw,kw,kf.

15. or/12-14

16. 4 and 11 and 15

17. limit 16 to yr="2021 -Current"

Fields: hw ([subject] heading word); kf (keyword heading word (MEDLINE)); kw (keyword (Embase); keyword heading (MEDLINE)); tw (text word).

**Web of Science** (from 30 June 2021 to 2025 January 24)

(TI=((("Mendelian randomization" or "Mendelian randomisation" or "genetic instrument*" or ("genetic variant*" and "instrumental variable*")) AND (scale* or checklist* or “critical apprais*” or tool* or guide* or dictionary or glossary) AND (valid* or quality or ((bias or confound*) and (asses* or measur* or evaluat*)))))) OR

(AB=((("Mendelian randomization" or "Mendelian randomisation" or "genetic instrument*" or ("genetic variant*" and "instrumental variable*")) AND (scale* or checklist* or “critical apprais*” or tool* or guide* or dictionary or glossary) AND (valid* or quality or ((bias or confound*) and (asses* or measur* or evaluat*)))))) OR (AK=((("Mendelian randomization" or "Mendelian randomisation" or "genetic instrument*" or ("genetic variant*" and "instrumental variable*")) AND (scale* or checklist* or “critical apprais*” or tool* or guide* or dictionary or glossary) AND (valid* or quality or ((bias or confound*) and (asses* or measur* or evaluat*))))))

Fields: TI: title; AB: abstract; AK: author keyword;

**bioRxiv and medRxiv**: Mendelian AND randomi*ation

**PROSPERO:** Mendelian randomization OR Mendelian randomisation

**Studies excluded at full text with exclusion reasons.**

**Background paper**

1. Spiga F, Gibson M, Dawson S, Tilling K, Davey Smith G, Munafo MR, Higgins JP. Tools for assessing quality and risk of bias in Mendelian randomization studies: a systematic review. International Journal of Epidemiology. 2023 Feb 1;52(1):227-49.
2. Kang H, Lee Y, Cai TT, Small DS. Two robust tools for inference about causal effects with invalid instruments. Biometrics. 2022 Mar;78(1):24-34.

**Duplicated and no tool (4 references)**

1. Levin MG, Burgess S. Mendelian randomization as a tool for cardiovascular research: a review. JAMA cardiology. 2024 Jan 1;9(1):79-89.
2. Levin MG, Burgess S. Mendelian randomization as a tool for cardiovascular research: a review. JAMA cardiology. 2024 Jan 1;9(1):79-89.
3. Tang Z, Can L, Xuan S, Chen L, Zhang J, Zhang B, Wan X, Li Z, Tang F, He Z. Unveiling the Etiology of Urological Tumors: A Systematic Review of Mendelian Randomization Applications in Renal Cell Carcinoma, Bladder Cancer, and Prostate Cancer. Urology Journal. 2024 May 2;21(05):283-92.
4. Tang Z, Can L, Xuan S, Chen L, Zhang J, Zhang B, Wan X, Li Z, Tang F, He Z. Unveiling the Etiology of Urological Tumors: A Systematic Review of Mendelian Randomization Applications in Renal Cell Carcinoma, Bladder Cancer, and Prostate Cancer. Urology Journal. 2024 May 2;21(05):283-92.

**No newly developed tool other than STROBE-MR checklist provided (15 references)**

1. Ho J, Mak CC, Sharma V, To K, Khan W. Mendelian randomization studies of lifestyle-related risk factors for osteoarthritis: a PRISMA review and meta-analysis. International journal of molecular sciences. 2022 Oct 7;23(19):11906.
2. Ibrahim M, Thanigaimani S, Singh TP, Morris D, Golledge J. Systematic review and Meta-Analysis of Mendelian randomisation analyses of Abdominal aortic aneurysms. IJC Heart & Vasculature. 2021 Aug 1;35:100836.
3. Li D, Hong X, Chen T. Association between rheumatoid arthritis and risk of parkinson's disease: A meta-analysis and systematic review. Frontiers in Neurology. 2022 May 11;13:885179.
4. Zhang W, Ghosh D. A general approach to sensitivity analysis for Mendelian randomization. Statistics in biosciences. 2021 Apr;13(1):34-55.
5. Burgess S, Mason AM, Grant AJ, Slob EA, Gkatzionis A, Zuber V, Patel A, Tian H, Liu C, Haynes WG, Hovingh GK. Using genetic association data to guide drug discovery and development: Review of methods and applications. The American Journal of Human Genetics. 2023 Feb 2;110(2):195-214.
6. Saccaro LF, Gasparini S, Rutigliano G. Applications of Mendelian randomization in psychiatry: a comprehensive systematic review. Psychiatric Genetics. 2022 Dec 1;32(6):199-213.
7. Julian TH, Boddy S, Islam M, Kurz J, Whittaker KJ, Moll T, Harvey C, Zhang S, Snyder MP, McDermott C, Cooper-Knock J. A review of Mendelian randomization in amyotrophic lateral sclerosis. Brain. 2022 Mar 1;145(3):832-42.
8. Dai X, Wang H, Zhong R, Li J, Hou Y. Causality of genetically determined metabolites on susceptibility to prevalent urological cancers: a two-sample Mendelian randomization study and meta-analysis. Frontiers in Genetics. 2024 Jul 1;15:1398165.
9. Cara KC, Taylor SF, Alhmly HF, Wallace TC. A Systematic Review and Meta-Analysis of Vitamin D Intakes and Status in Adults With Irritable Bowel Syndrome. Current Developments in Nutrition. 2024 Jul 1;8.
10. Wei J, Zhu X, Wang J, Yang K, Chen J, Li J, Zuo S, Liu N. Mendelian randomization studies in ankylosing spondylitis: a systematic review. International Journal of Rheumatic Diseases. 2024 Nov;27(11):e15408.
11. Lovegrove CE, Howles SA, Furniss D, Holmes MV. Causal inference in health and disease: a review of the principles and applications of Mendelian randomization. Journal of Bone and Mineral Research. 2024 Nov;39(11):1539-52.
12. Sun Y, Liu Y, Dian Y, Zeng F, Deng G, Lei S. Association of glucagon-like peptide-1 receptor agonists with risk of cancers-evidence from a drug target Mendelian randomization and clinical trials. International Journal of Surgery. 2024 Aug 1;110(8):4688-94.
13. Guan J, Liu T, Gao G, Yang K, Liang H. Associations between lifestyle-related risk factors and back pain: a systematic review and meta-analysis of Mendelian randomization studies. BMC Musculoskeletal Disorders. 2024 Aug 1;25(1):612.
14. Fang A, Zhao Y, Yang P, Zhang X, Giovannucci EL. Vitamin D and human health: evidence from Mendelian randomization studies. European journal of epidemiology. 2024 May;39(5):467-90.
15. Mikkelsen H, Landt EM, Benn M, Nordestgaard BG, Dahl M. Causal risk factors for asthma in Mendelian randomization studies: A systematic review and meta‐analysis. Clinical and translational allergy. 2022 Nov;12(11):e12207.
